# Comparisons of COVID-19 dynamics in the different countries of the World using Time-Series clustering

**DOI:** 10.1101/2020.08.18.20177261

**Authors:** Emiliano Alvarez, Juan Gabriel Brida, Erick Limas

## Abstract

In recent months, the world has suffered from the appearance of a new strain of coronavirus, causing the COVID-19 pandemic. There are great scientific efforts to find new treatments and vaccines, at the same time that governments, companies, and individuals have taken a series of actions in response to this pandemic. These efforts seek to decrease the speed of propagation, although with significant social and economic costs. Countries have taken different actions, also with different results. In this article we use non-parametric techniques (HT and MST) with the aim of identifying groups of countries with a similar spread of the coronavirus. The variable of interest is the number of daily infections per country. Results show that there are groups of countries with differentiated contagion dynamics, both in the number of contagions plus at the time of the greatest transmission of the disease. It is concluded that the actions taken by the countries, the speed at which they were taken and the number of tests carried out may explain part of the differences in the dynamics of contagion.

## 1 Introduction

In recent months, the world has suffered from the appearance of a new strain of coronavirus, causing the COVID-19 pandemic. There are great scientific efforts to find new treatments and vaccines, at the same time that governments, companies, and individuals have taken a series of actions in response to this pandemic. These efforts seek to decrease the speed of propagation, although with significant social and economic costs. Countries have taken different actions, also with different results. In this article non-parametric techniques (HT and MST) are introduced to identify groups of countries with a similar spread of the coronavirus evolution. The variable of interest is the number of daily infections per country during a period of at least 100 days after the confirmation of the tenth case. The coronavirus propagation is analyzed in terms of a complex system composed of many interacting elements that exhibit numerous forms of emergent collective dynamics (Machado and Lopes, 2020). Complex systems cannot be explained by studying the constituent parts in isolation, but by considering the interrelation with the other elements of this system.

Vasconcelos et al. (2020) analyze the fatality curves of the COVID-19 disease, represented by the cumulative number of deaths, for eight countries: Brazil China, France, Germany, Iran, Italy, South Korea, and Spain. The study model the cumulative number of deaths by applying the Richards growth model and find that it describes very well the fatality curves in all cases. The authors also find that-in order to be effective-the countermeasures (i.e. social distancing, quarantine, school closures, etc.) must be taken immediately after the onset of the epidemic. In particular, they find that if the interventions are delayed by ten days the respective efficiencies drop to about 50% or less. Additionally, a delay of additional 20 days brings the efficiency to less than 30%.

Manchein et al. (2020) study the growth of the cumulative number of confirmed infected cases by a novel coronavirus (COVID-19) until March 27, 2020. The study focuses on nine countries (Brazil, China, France, Germany, Italy, Japan, Spain, Republic of Korea, and USA) and finds that in all cases the cumulative number of confirmed infected cases follows a power-law growth. Moreover, by using the distance correlation, the power-law curves between countries are statistically highly correlated. The authors argue that these correlations suggest the universality of such curves around the world. Regarding the impact of the countermeasures, the article finds that soft quarantine strategies are inefficient to flatten the growth curves. Among the most effective countermeasures to flatten the power laws, the authors identify the social distancing of individuals and also the strategy of identifying and isolating infected individuals which, according to the authors, is the best strategy to flatten the curves.

Zarikas et al. (2020) present a hierarchical cluster analysis, in which they analyze active cases, active cases per population and active cases per population and per area based on Johns Hopkins epidemiological data. The analysis identifies four different shapes in the evolution rate of COVID-19. The first shape corresponds to China, which characterizes by a sharp increase during the first 21 days and then a flattening of the curve2. The second shape corresponds to South Korea, which is similar to the China’s shape but with a lower number of cases. The third shape is associated to the countries where the cases appeared early, but they have a low number of cases (for example, Vietnam, Thailand, Japan, Japan, Singapore or Nepal). Finally, the fourth shape corresponds to countries in which the cases arose since several days, but the sharp increase occurred very recently and the number of cases during last day was very high (for example, USA, France, Germany, Italy, UK, Spain, Iran, Canada or Israel. Important to note, as the authors suggest, that the surface area of each country is a parameter influencing the criticality of the situation, i.e. geography matters.

Chandu (2020) utilizes a K-means clustering algorithm to analyze 89 countries with at least 1000 confirmed positive cases. The algorithm groups the countries into two main clusters. The Americas, European countries, and Australia were members of the same cluster, which is characterized by high COVID-19 case fatality rate, higher proportion of country’s population tested COVID-19 positive, higher percentage of GDP spent as public health expenditure, and greater percentage of population being more than 65 years of age.

Rojas et al. (2020) present an analysis of the states of the United States with the aim of measuring the similarity of COVID-19 time series. In the proposed methodology, infected and deceased persona are jointly analyze and the similarity between time series is measured through the dynamic time warping distance (an extension of the Euclidean distance) and nine different clusters are obtained, showing a different pattern in the eastern region and western region of the United States.

In this article we use non-parametric techniques (HT and MST) with the aim of identifying groups of countries with a similar spread of the coronavirus. The variable of interest is the number of daily infections per country. The remainder of this paper is organized as follows. In section 2, the methodology of this paper is presented: Hierarchical Trees (HT) and the Minimum Spanning Trees (MST). Section 3 introduces the data, whereas section 4 presents the empirical results. The final section 5 draws the conclusions.

## 2 Methodology

Following the methodology developed by Mantegna (1999), in this study the coronavirus propagation is formulated as a network problem, where each country would be represented as a node, and the relationship between each pair of countries as a link. Thus, the spread of coronavirus would be a complex network of mutually interacting nodes and each link would represent how similar is the coronavirus dynamics between any pair of countries.

The Minimal Spanning Tree (MST) graphs were introduced by Kruskal in graph theory (Kruskal, 1956), but they started to be utilized in the realm of Econophysics due to the work of Mantegna (1999), who utilized the MST applied to time series data to analyze stock market correlations. The clustering process used to form the MST is known in cluster analysis as the single-linkage clustering method. The main advantage of the MST is that it simplifies comparisons by reducing the number of elements that must be analyzed. Moreover, since MST selects the strongest relationships among network nodes, it condenses the core information on the global structure of the network. Thus, the general characteristics of the network can be reproduced in the MST (Hill, 1999; Kwapień et al., 2009). In this article we propose analyzing the evolution of new daily coronavirus cases by using elements of graph theory. Thus, the time series of new cases per country will be the nodes of a graph, while the relationships between countries will be represented by the links of the graph. This way, the links correspond to the Pearson’s correlation distance (Mantegna, 1999) between any pair of countries. Following Stanley and Mantegna (2000), from these correlation distance we construct the minimum spanning tree (MST), which is the minimal graph that covers all nodes without loops. The first step of this methodology is the calculation of the Pearson’s correlations between the n × n pairs of countries:

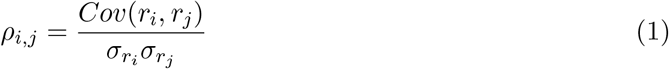

where *r_i_* is the number of new cases in country *i*. Then, the correlation matrix is built with the correlation coefficients *ρ_i,j_*. By definition, *ρ_i,j_* takes values in the interval (−1, 1), where −1 means complete anti-correlation, 1 complete correlation and 0 that the two variables are uncorrelated. This matrix is symmetrical, with *ρ_i,j_* = 1 in this main diagonal. As it is well known, the Pearson correlation coefficient (1) does not fulfill the three axioms that define a Euclidean metric. For this reason, the correlation matrix is transformed into the correlation distance matrix according que the following formula

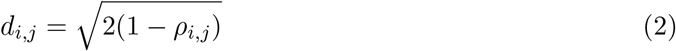

which fulfills the three axioms of an Euclidean distance:

i. *d*(*i, j*) = 0 *↔ i* = *j*
ii. *d*(*i, j*) = *d*(*j, i*)
iii. *d*(*i, j*) *≤ d*(*i, k*) + *d*(*k, j*)

Then, the distance matrix is used to determine the minimal spanning tree (MST) connecting the *n* vertices. The MST is constructed following the Kruskal’s algorithm, which links all the vertices together in a single graph that minimizes the distances between the corresponding time series. Then, the distance matrix is used to determine the minimal spanning tree (MST) connecting the *n* vertices. The Kruskal method is implemented by following the next steps. First, the algorithm chooses a pair of nodes with the nearest distance and connects them with a line that is proportional to the distance. Then, the algorithm connects another pair of nodes with the second nearest distance. In the third step, the nearest pair that is not connected by the same tree is also linked. This step is repeated until all the nodes are connected in a single tree.

From the MST we obtain the subdominant ultrametric distance matrix X, which corresponds to the matrix whose elements are the subdominant ultrametric distance *d^∗^*(*i, j*). The subdominant ultrametric distance *d^∗^*(*i, j*) between the vertices *i* and *j* is the maximum value of any correlation distance *d*(*i, j*) detected by moving one step from *i* to *j* through the shortest path connecting *i* and *j* in the MST. The ultrametric distance *d^∗^*(*i, j*) between the vertices *i* and *j* is then given by:

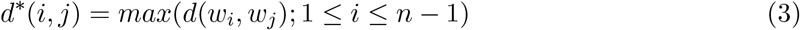

where ((*w*_1_*, w*_2_), (*w*_3_*, w*_4_),…, (*w_n−_*_1_*, w_n_*)) shows the minimal path in the MST that connects the vertices *i* and *j*, where *w*_1_ = 1 and *w_n_* = *j*. Using this formula we compute the ultrametric distance *d^∗^*(*i, j*) between each pair of currencies and from it we construct the Hierarchical Tree (HT), which is represented as a tree known as dendogram.

## 3 Data

This study compares the active cases per population of 191 countries. For each country, we construct a time series composed by daily data of the number of daily infections per country. Each time series considers a period of 100 days after the confirmation of the tenth case, using a moving average smoothing. This way, the analysis of new active cases takes into account the different moments in which the virus began to disseminate in each country. Data are obtained from “Our World in Data” (Roser et al., 2020), which is built from information supplied by the European Centre for Disease Prevention and Control, government reports, Oxford COVID-19 Government Response Tracker, World Bank – World Development Indicators, United Nations Statistics Division and Eurostat. Although this study focuses on new active cases per population, it will also analyze total active cases, new deaths and total deaths per population, new tests and test per 1,000 people, the Government Response Stringency Index as well as other demographic and socioeconomic indicators.

## 4 Results

In this section we present the empirical results of this article. Figure 1 shows the MST and the corresponding HT. In Appendix 1, Table 2 lists the countries belonging to each cluster and Figure 8 shows how the clusters are distributed among the different countries belonging to the sample.

According to pesudo-F (Caliński and Harabasz, 1974) and pseudo-t2 (Duda et al., 1973) hierarchical clustering stopping rules, three main groups can be identified plus a fourth small group integrated by both Mongolia and the series representing the average of all the countries. Cluster 1 has 104 members, cluster 2 is integrated by 43 countries and cluster 3 includes 43 countries. Within these groups represented in Figure 1, it can be seen that the structure of the network for cluster 2 is linear while for clusters 1 and 3 it has countries with a higher number of connections. A higher number of connections (higher node degree) or a higher number of shortest paths that pass through the node (higher betweenness centrality) imply that these nodes summarize a large amount of information from the network. In other words, it means the data series for these countries represent many of the characteristics of the groups to which they belong. Table 1 shows that the nodes with the highest centrality are India, Indonesia and Mexico for cluster 1 and Italy and Romania for cluster 3.

**Table 1:** Node degree and node betweenness for the set of countries with the highest betweenness centrality.

| Country | Group | Node degree | Node betweenness |
| --- | --- | --- | --- |
| India | 1 | 7 | 11486 |
| Italy | 3 | 5 | 9783 |
| Indonesia | 1 | 4 | 9659 |
| Mexico | 1 | 4 | 9398 |
| Romania | 3 | 5 | 9339 |

**Figure 1:**
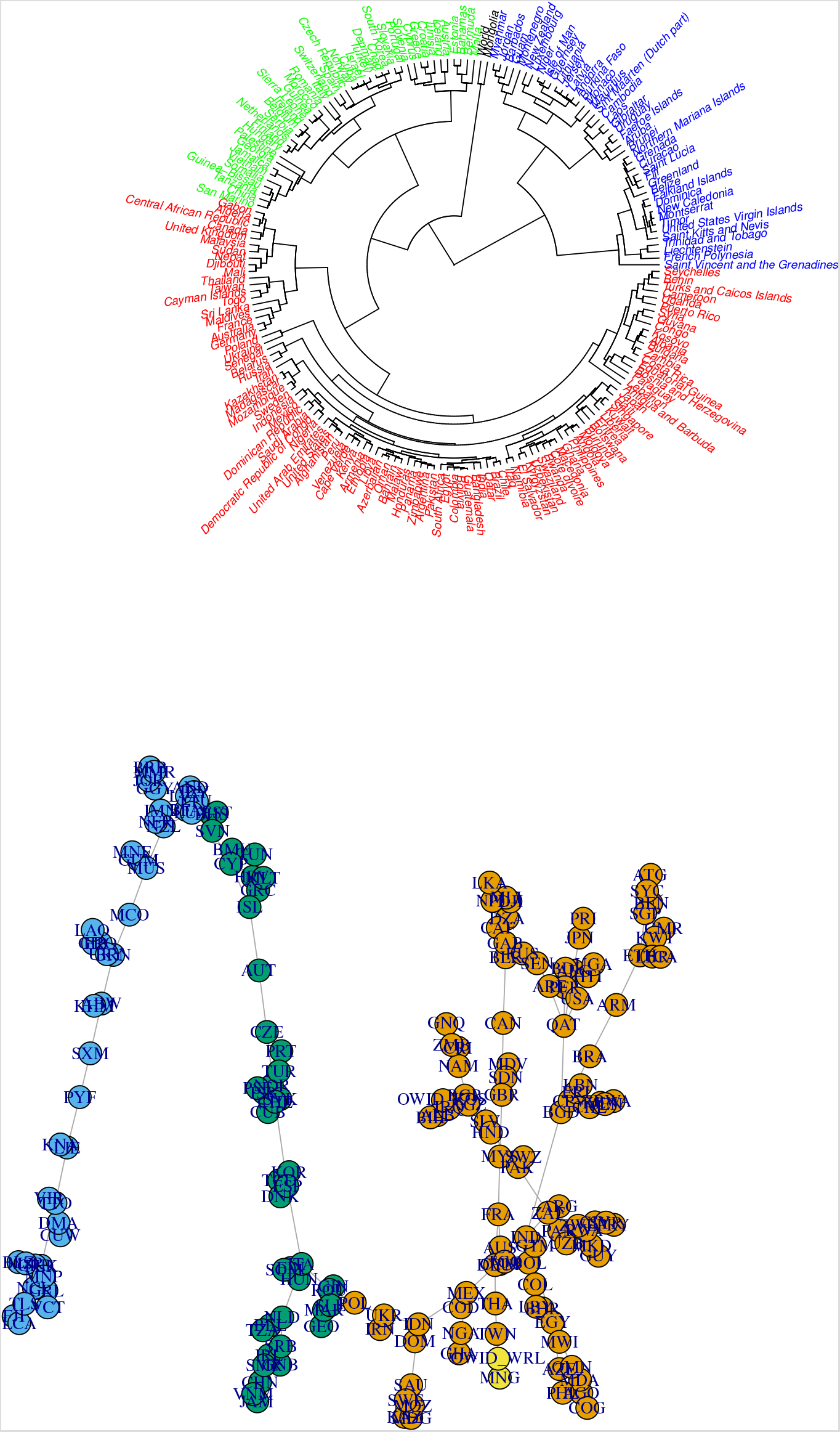
Hierarchical Tree (up) and Minimal Spanning Tree (down)

As showed by Figure 2, each group of countries exhibits a different kind of dynamic behavior. Countries in cluster 1 starts with a low number of cases per million, although it maintains an upward trend during the whole period. On the other hand, cluster 2 begins with values between 10 and 20 cases per million and follows a decreasing trend. Finally, cluster 3 reaches its peak between the 20th and 40th day and immediately starts to decrease. On the other hand, when observing the dynamics within each group of countries, it can be seen that the median represents the trend behavior within each group. How similarity is measured in this work (Pearson’s correlation coefficient) tends to group countries with similar trends, therefore the three main clusters represent three different dynamic behaviors for the first 100 days: sustained growth, decrease sustained or accelerated growth followed by a decrease in the number of new cases.

**Figure 2:**
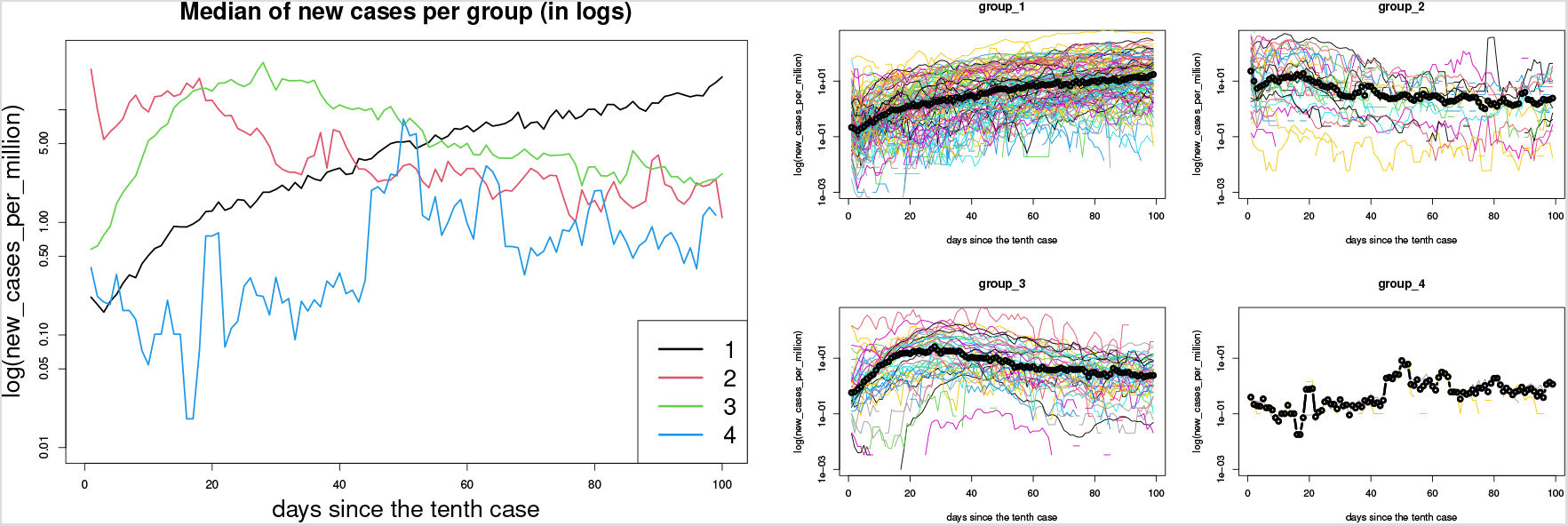
Evolution of new cases: median of each group, in logs (left) and joint graph of the elements of each cluster, in logs (right).

Figure 3 shows examples of the dynamic behavior of countries extracted from each cluster. Mexico and Argentina, both members of cluster 1, follow a quite similar upward trend, whereas the paradigmatic case of Uruguay (see Moreno et al, 2020)-member of cluster 2-follows an oscillatory although descending trend. Italy is a representative example of cluster 3, as we mentioned before. We observe that it began with a pronounced increase in new cases, reached its peak between the 20th and 40th day and afterwards started a descending trajectory.

**Figure 3:**
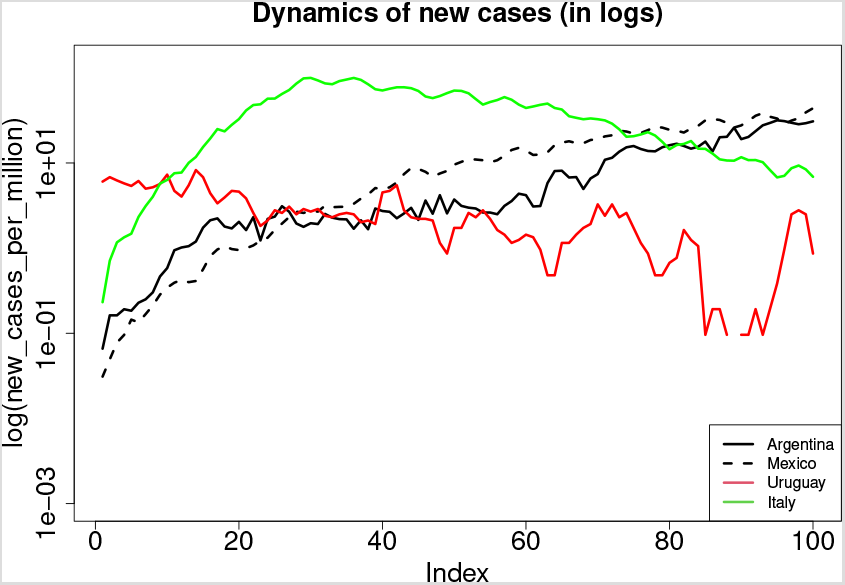
Dynamics of new cases

It is worth to note that cluster 3 is the group with the highest number of cases, as can be seen in Figure 4. In addition, this cluster has had the most pronounced downward trend. This group includes China and Italy, countries with an accelerated initial growth. Initially they experienced an accelerated rate of contagion, reached their respective peaks and entered sooner into the phase of deceleration. In addition, many of the cluster 3 countries have been the first countries to face COVID 19, in the epidemic stage. We can assume that in the absence of knowledge about this disease and the lack of adequate protocols in early 2020, the virus spread more rapidly through these countries. Group 2 is composed of islands and small countries. We can assume that these geographical conditions made it easier for them to isolate themselves once the contagion stage began, preventing the possibility of new infections from people who travel from abroad.

**Figure 4:**
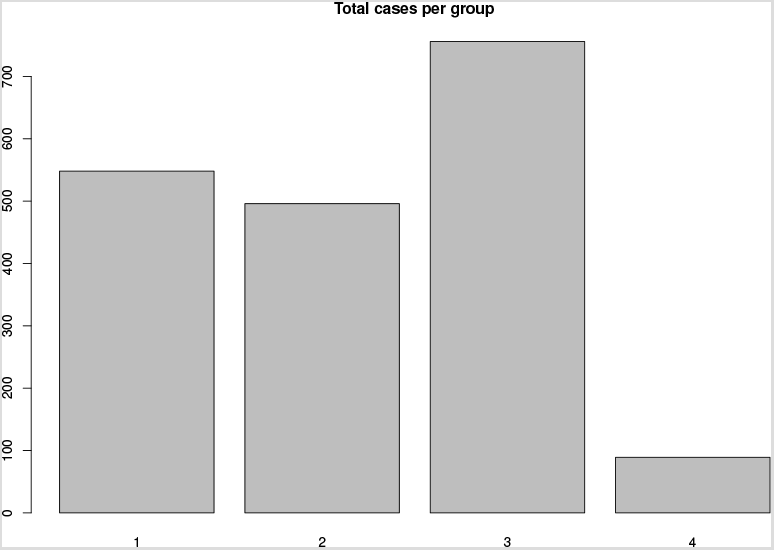
Total number of infections in the period by group (median of each group).

Total number of deaths and new tests per thousand inhabitants are shown in Figure 5. We observe that groups 2 and 3 began to stabilize between the 40th and 60th day, whereas the cluster 1 has maintained a constant growth rate since day 30. With respect to new tests, group 2 is the one with the highest number, followed by cluster 3. Group 1 registers a lower number of new tests, although with an increasing trend. An interesting result is that the groups of countries that have achieved a drop in the number of cases are those that perform a greater number of tests; moreover, it after 100 days, with a growing trend and a higher number of new cases per million, the countries of cluster 1 are maintained below the number of tests performed. This may mean that, along with other measures, mass testing helps control the spread^1^.

Figure 6 presents a comparison between groups according to demographic and socioeconomic indicators. From the comparison, we observe that group 3 shows better socioeconomic indicators than the other groups. However it also present higher risk factors due to the age of its population. The former might be related to a greater effectiveness in the effective control of disease, whereas the latter to a higher mortality rates. On the other hand, group 1 has, in median, the largest population, the lowest density, younger population and worse socioeconomic conditions. For most of the indicators, group 2 occupies the intermediate position, except for its population and the proportion of the population with access to basic handwashing facilities.

**Figure 5:**
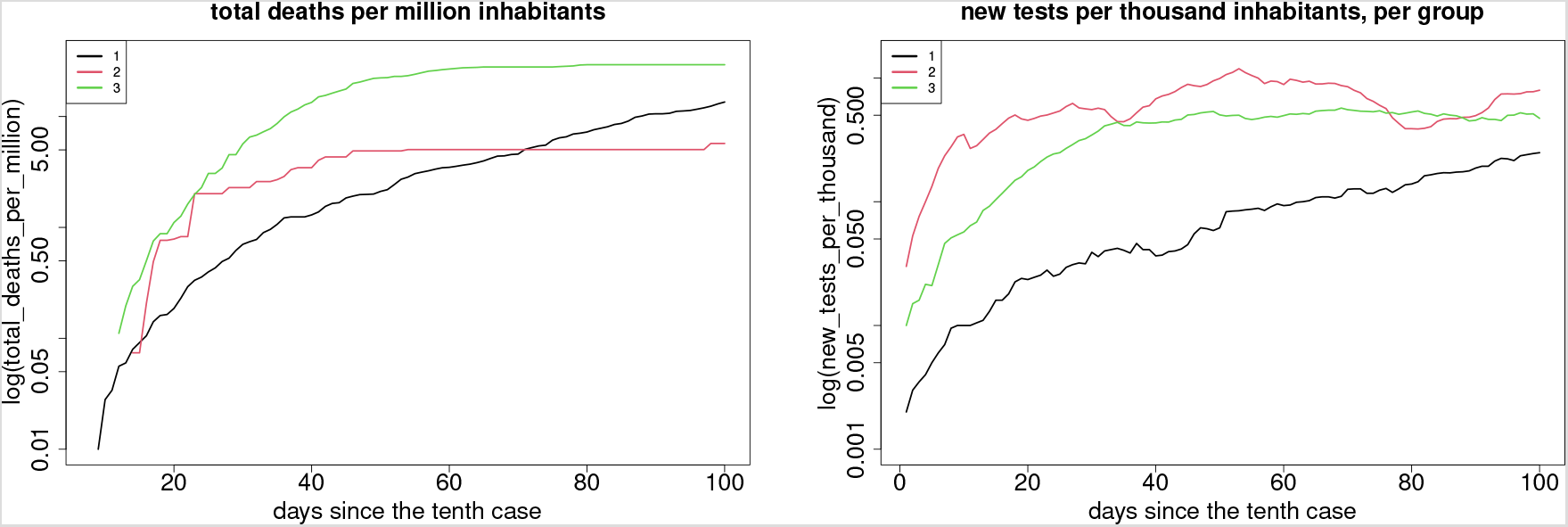
Total deaths per million inhabitants (left) and new tests per thousand inhabitants (right).

**Figure 6:**
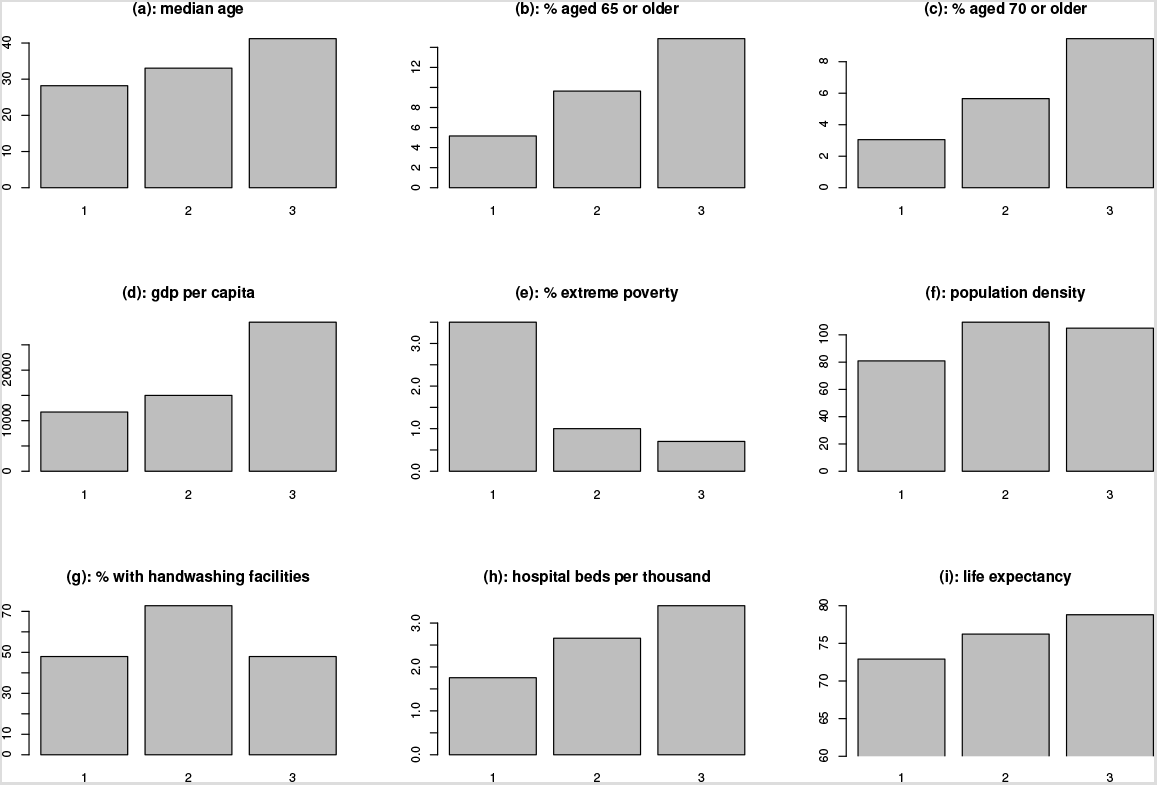
Demographic and socioeconomic indicators.

Finally, Figure 7 presents the Stringency Index, which is a composite measure based on nine response indicators, including school closures, workplace closures, and travel bans, rescaled to a value from 0 to 100, where 100 represents the strictest response. The groups reach similar levels of restrictions about 20 days after the first cases, although relevant differences are observed. Group 1 progressively increases their controls, which began to stabilize around day 50. Group 2 had the quickest response, reaching its peak before the day 20. Group 3 applied the measures more slowly, following the contagion curve. Relationships between the Stringency index and data about new cases per million can be established for each group, observing that those countries with a faster government response have been more successful in controlling the disease, while a decrease in the number of new infections does not seem to be related to mobility restrictions more extended in time. An example of this is cluster 1, which has remained at very high levels of restrictions from the 20th to the end of the period analyzed without being able to modify the growing trend of new infections. Countries in clusters 2 and 3 have progressively reduced restrictions on mobility, following the decrease in new cases in those clusters.

**Figure 7:**
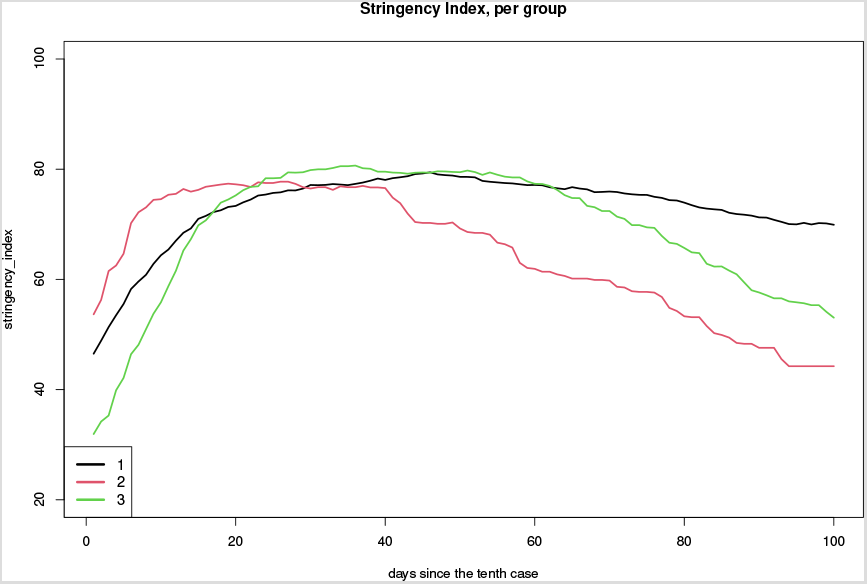
Stringency index.

## 5 Conclusions

Three large clusters of countries were identified and each one was associated with three different types of contagion curves. The largest cluster has not reached its maximum yet and maintains a growing trend. It is also noteworthy that cluster 1 has higher levels of poverty, which implies that a significant proportion of the population must seek their livelihood on a daily basis, also related to the lesser scope of their health and social protection systems, in accordance with Goutte et al. (2020). For this segment of the population staying at home during the pandemic is a major challenge. This might be one the drivers behind the fact that in cluster 1 the virus continues spreading. This dynamics can be seen also in developed countries with weakened organized labor and job insecurity, because as McKee and Stuckler (2020) notes, “even the wealthiest societies are only as strong as their weakest members”. The cluster 2 was the first to reach the peak of daily infections and quickly entered into phase of decline. It is worth observing that this group is composed by small countries and islands, which highlights the importance of geography as a key factor (Zarikas et al., 2020). The cluster 3 had an abrupt increase in new cases and once the maximum was reached it entered into a decreasing phase. This cluster also had both the highest death toll and the oldest age group, which underlines the importance of protecting people in risk factor groups. Another important point is the relationship between the Stringency index and the contagious curves. We observed that the three groups reached similar levels of restrictions 20 days after their first cases. However, cluster 2 implemented countermeasures 10 days before the other clusters and reached its maximum without experiencing an abrupt jump in the number of cases. This result highlights the importance of a timely response and it is in line with Vasconcelos et al. (2020), in which the authors find that the countermeasures must be taken immediately after the onset of the epidemic. The quality of the health and social protection systems are crucial in this regard since both the level and the speed of the government response will depend on these factors, as can be seen also in Armocida et al. (2020). The levels of trust of citizens in science and institutions and the association between academia and political representatives (see Moreno et al. (2020)) are essential for good management of the situation. An additional implication on this issue is about the dichotomy between the economy and health. Countries of clusters 2 and 3, which have been more aggressive and faster in implementing stay-at-home measures and massive testing, have achieved a decrease in the number of new cases and a progressive decrease in restrictions on the mobility, while cluster 1 countries maintain higher restrictions on mobility after one hundred days. Lin and Meissner (2020) do not find an association between local restriction policies and the economy, although unlike what Lin and Meissner (2020) suggested, our analysis suggest that these measures could be associated with changes in the number of infections. It is necessary to investigate more about this matter, but it could be thought that there is no dichotomy between health and the economy, but that the best measures for subsequent reactivation of economic activity consist of the rapid response to the pandemic. This aid should cover both health systems and the most vulnerable individuals in society. One important limitation when working with these databases is the possibility of under-reporting of cases, which might imply-among other consequences-that cluster 1 has already reached its maximum or that cluster 2 had a less/more pronounced peak. Additionally, it is important to note that at this point it is not clear if all the reported deaths are caused by COVID-19 or if the deaths were caused by a third factor. This might constitute a future research question. In addition, this work may be extended in order to analyze other variables, such as the ratios “deaths / active-cases” and “active-cases / tests” or by utilizing temporal windows to study the clusters dynamics.

## Data Availability

The data used in this manuscript are available at https://ourworldindata.org/coronavirus

## Appendix

**Table 2:** Countries belonging to each cluster.

| Group 1 |  |  |  |
| --- | --- | --- | --- |
| Afghanistan | Congo | Kosovo | Qatar |
| Albania | Costa Rica | Kuwait | Russia |
| Algeria | Cote d'Ivoire | Kyrgyzstan | Rwanda |
| Angola | Democratic Rep. of Congo | Lebanon | Saudi Arabia |
| Antigua and Barbuda | Djibouti | Liberia | Senegal |
| Argentina | Dominican Republic | Libya | Seychelles |
| Armenia | Egypt | Macedonia | Singapore |
| Australia | El Salvador | Madagascar | South Africa |
| Azerbaijan | Equatorial Guinea | Malawi | Sri Lanka |
| Bahrain | Eritrea | Malaysia | Sudan |
| Bangladesh | Ethiopia | Maldives | Swaziland |
| Belarus | France | Mali | Sweden |
| Benin | Gabon | Mexico | Syria |
| Bolivia | Germany | Moldova | Taiwan |
| Bosnia and Herzegovina | Ghana | Mozambique | Thailand |
| Botswana | Guatemala | Namibia | Togo |
| Brazil | Guyana | Nepal | Turks and Caicos Islands |
| Bulgaria | Haiti | Nigeria | Uganda |
| Burundi | Honduras | Oman | Ukraine |
| Cameroon | India | Pakistan | United Arab Emirates |
| Canada | Indonesia | Panama | United Kingdom |
| Cape Verde | Iran | Paraguay | United States |
| Cayman Islands | Iraq | Peru | Uzbekistan |
| Central African Republic | Japan | Philippines | Venezuela |
| Chile | Kazakhstan | Poland | Zambia |
| Colombia | Kenya | Puerto Rico | Zimbabwe |
| Group 2 |  |  |  |
| Andorra | Fiji | Latvia | Niger |
| Aruba | French Polynesia | Liechtenstein | Northern Mariana Islands |
| Barbados | Gibraltar | Lithuania | Saint Kitts and Nevis |
| Belize | Greenland | Luxembourg | Saint Lucia |
| Brunei | Grenada | Mauritius | Saint Vincent and the Grenadines |
| Burkina Faso | Guam | Monaco | Sint Maarten |
| Cambodia | Guernsey | Montenegro | Timor |
| Curacao | Isle of Man | Montserrat | Trinidad and Tobago |
| Dominica | Jersey | Myanmar | United States Virgin Islands |
| Faeroe Islands | Jordan | New Caledonia | Uruguay |
| Falkland Islands | Laos | New Zealand |  |
| Group 3 |  |  |  |
| Austria | Estonia | Jamaica | Slovakia |
| Bahamas | Finland | Malta | Slovenia |
| Belgium | Georgia | Morocco | Somalia |
| Bermuda | Greece | Netherlands | South Korea |
| Chad | Guinea | Norway | Spain |
| China | Guinea-Bissau | Palestine | Switzerland |
| Croatia | Hungary | Portugal | Tanzania |
| Cuba | Iceland | Romania | Tunisia |
| Cyprus | Ireland | San Marino | Turkey |
| Czech Republic | Israel | Serbia | Vietnam |
| Denmark | Italy | Sierra Leone |  |
| Group 4 |  |  |  |
| Mongolia | World |  |  |

Figure 8 shows the geographical distribution of the clusters

**Figure 8:**
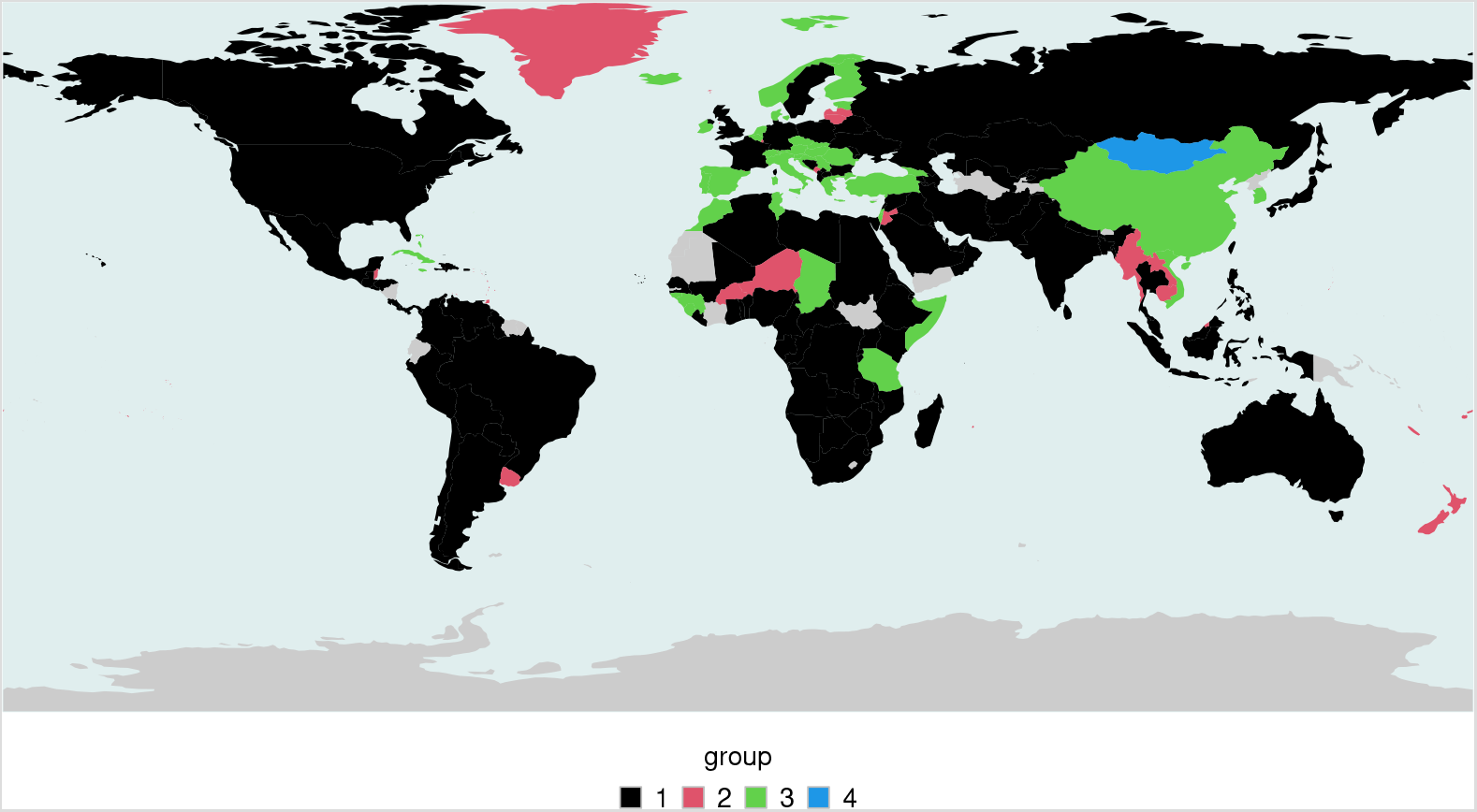
World map with geographic distribution of clusters, found from hierarchical clustering.

## Footnotes

1 The intensity of the tests is a variable of government policy that is related to the success in containing the disease. This association can also be found in James and Menzies (2020).

## Notes

### Competing Interest Statement

The authors have declared no competing interest.

### Funding Statement

No external funding was received

### Author Declarations

All relevant ethical guidelines have been followed.

